# Safety and exceptional immunogenicity of novel 5T4 viral vectored vaccination regimes in early stage prostate cancer: a phase I clinical trial

**DOI:** 10.1101/2020.03.05.20031500

**Authors:** Federica Cappuccini, Richard Bryant, Emily Pollock, Lucy Carter, Clare Verrill, Julianne Hollidge, Ian Poulton, Megan Baker, Celia Mitton, Andrea Baines, Armin Meier, Guenter Schmidt, Richard Harrop, Andrew Protheroe, Ruth MacPherson, Steven Kennish, Susan Morgan, Selena Vigano, Pedro Romero, Thomas Evans, James W.F. Catto, Freddie Hamdy, Adrian V.S. Hill, Irina Redchenko

**Affiliations:** The Jenner Institute, University of Oxford, Oxford, UK; Nuffield Department of Surgical Sciences, University of Oxford, Oxford, UK; Department of Urology, Churchill Hospital Oxford University Hospitals NHS Foundation Trust, Oxford, UK; Oxford NIHR Biomedical Research Centre, University of Oxford, Oxford, UK; Definiens GmbH, Munich, Germany; Oxford Biomedica UK limited, Oxford, UK; Department of Oncology, Oxford Cancer and Haematology Centre, Churchill Hospital Oxford University Hospitals NHS Foundation Trust, Oxford, UK; Department of Radiology, Churchill Hospital Oxford University Hospitals NHS Foundation Trust, Oxford, UK; Department of Radiology, Sheffield Teaching Hospitals NHS Trust, Sheffield, UK; Department of Pathology, Sheffield Teaching Hospitals NHS Trust, Sheffield, UK; Lausanne University Hospital, Lausanne, Switzerland; Vaccitech UK limited, Oxford, UK; Academic Urology Unit, University of Sheffield, Sheffield, UK

## Abstract

Prostate cancer (PCa) has been under investigation as a target for antigen-specific immunotherapies in metastatic disease settings for a decade. However, neither of the two clinically most developed prostate cancer vaccines, Sipuleucel-T and ProstVac, induce strong T cell immunity. In this first-in-man study, VANCE, we evaluated a novel vaccination platform based on two replication-deficient viruses, chimpanzee adenovirus (ChAd) and MVA (Modified Vaccinia Ankara), targeting the oncofetal self-antigen 5T4 in early stage PCa. Forty patients, either newly diagnosed with early stage prostate cancer and scheduled for radical prostatectomy or patients with stable disease on an active surveillance protocol, were recruited to the study to assess the vaccine safety and T cell immunogenicity. Secondary and exploratory endpoints included immune infiltration into the prostate, prostate specific antigen (PSA) change and assessment of phenotype and functionality of antigen-specific T cells. The vaccine had an excellent safety profile. Vaccination-induced 5T4-specific T cell responses were measured in blood by *ex vivo* IFN-γ ELISpot and were detected in the majority of patients with a mean level in responders of 198 spot-forming cells (SFC) per million peripheral blood mononuclear cells (PBMCs). Flow cytometry analysis demonstrated the presence of both CD8+ and CD4+ polyfunctional 5T4-specific T cells in the circulation. 5T4-reactive tumour infiltrating lymphocytes (TILs) were isolated from post-treatment prostate tissue. Some of the patients had a transient PSA rise 2-8 weeks following vaccination, possibly indicating an inflammatory response in the target organ. The potent T cell responses elicited support the evaluation of these vectored vaccine in efficacy trials.

## Introduction

PCa is the most common non-cutaneous malignancy in men and the second leading cause of male cancer-related death in the Western world (*1*). If the disease progresses to metastatic castration-resistant prostate cancer (mCRPC), the treatment options are mainly palliative in nature. Two next generation hormonal agents, abiraterone and enzalutamide, have improved treatment of mCRPC, however, resistance to these drugs eventually ensues, with the disease becoming lethal.

Cancer immunotherapy has been evolving over the last decade and may be a viable option for treatment of advanced stage prostate cancer patients. To date, the only therapeutic cancer vaccine to be approved by the US Food and Drug Administration (FDA) is Sipuleucel-T, and is indicated for patients with asymptomatic or minimally symptomatic mCRPC (*2*). This immunotherapy targets the prostate-specific antigen prostatic acid phosphatase (PAP) and is based on autologous peripheral-blood mononuclear cells (including antigen-presenting cells) activated *ex vivo* by cytokines and loaded with a recombinant fusion protein consisting of PAP and granulocyte-macrophage colony-stimulating factor (GM-CSF). The vaccine has shown modest, though statistically significant, efficacy in clinical trials, increasing overall survival in treated patients by four months. However, no effect on time to tumour progression compared to placebo group has been observed and the induced T cell responses appeared weak (*3*).

Another vaccine, ProstVac, has reached a phase III clinical trial (NCT01322490). This vaccine comprises two recombinant viral vectors, replication competent vaccinia and non-replicating fowlpox viruses, each encoding a transgene for PSA. In the phase II randomized placebo-controlled study in patients with metastatic prostate cancer men in the ProstVac group had a significant increase in median overall survival (8.5 months) compared to the control group. However, T cell immune responses against the vaccine-encoded antigen were very modest (*4*) and no clinical efficacy was observed in the phase III trial (*5*).

In this current study, we targeted the tumour antigen 5T4 (*6*) using an alternative immunisation platform based on a chimpanzee adenovirus (ChAd) prime and MVA boost (developed in our laboratories over a decade ago). This has been shown to be the most powerful approach for inducing polyfunctional protective T cell responses against antigens from diverse human pathogens in clinical trials (*7-13*). Preclinical evaluation of this vaccination strategy in murine prostate cancer models demonstrated its tumour protective efficacy (*14, 15*), thus providing a rationale for a phase I clinical study. Here we report the results of the first-in-human VANCE study, evaluating the safety of ChAdOx1-MVA 5T4 vaccine in early stage prostate cancer. The induction of antigen-specific immune responses in both the blood and the prostate gland, the effect of low dose cyclophosphamide (CTX) preconditioning on the immune responses and changes in serum PSA concentration are also reported. Vaccine efficacy assessments are effectively not possible in this trial design because the selected patient cohort either had a curative surgical treatment immediately after vaccination, or a subsequent very low risk of recurrent disease that would require a long follow-up period beyond the duration of this trial in order to assess any clinical impact of vaccination.

## Results

### Study participants and safety analysis

Forty patients were enrolled into the VANCE randomised open label phase I study between 26^th^ October 2015 and 19^th^ October 2017, with thirty-nine participants completing the study treatment and eligible for analysis (Fig. 1). All participants were diagnosed with low- or intermediate-risk prostate cancer scheduled for radical prostatectomy, or undergoing active surveillance for low-volume low- or intermediate-risk prostate cancer. Median age, clinical stage and Gleason score of trial participants are presented in Supplementary Table S1. VANCE clinical trial design is presented in Supplementary Fig. S1.

**Fig. 1.**
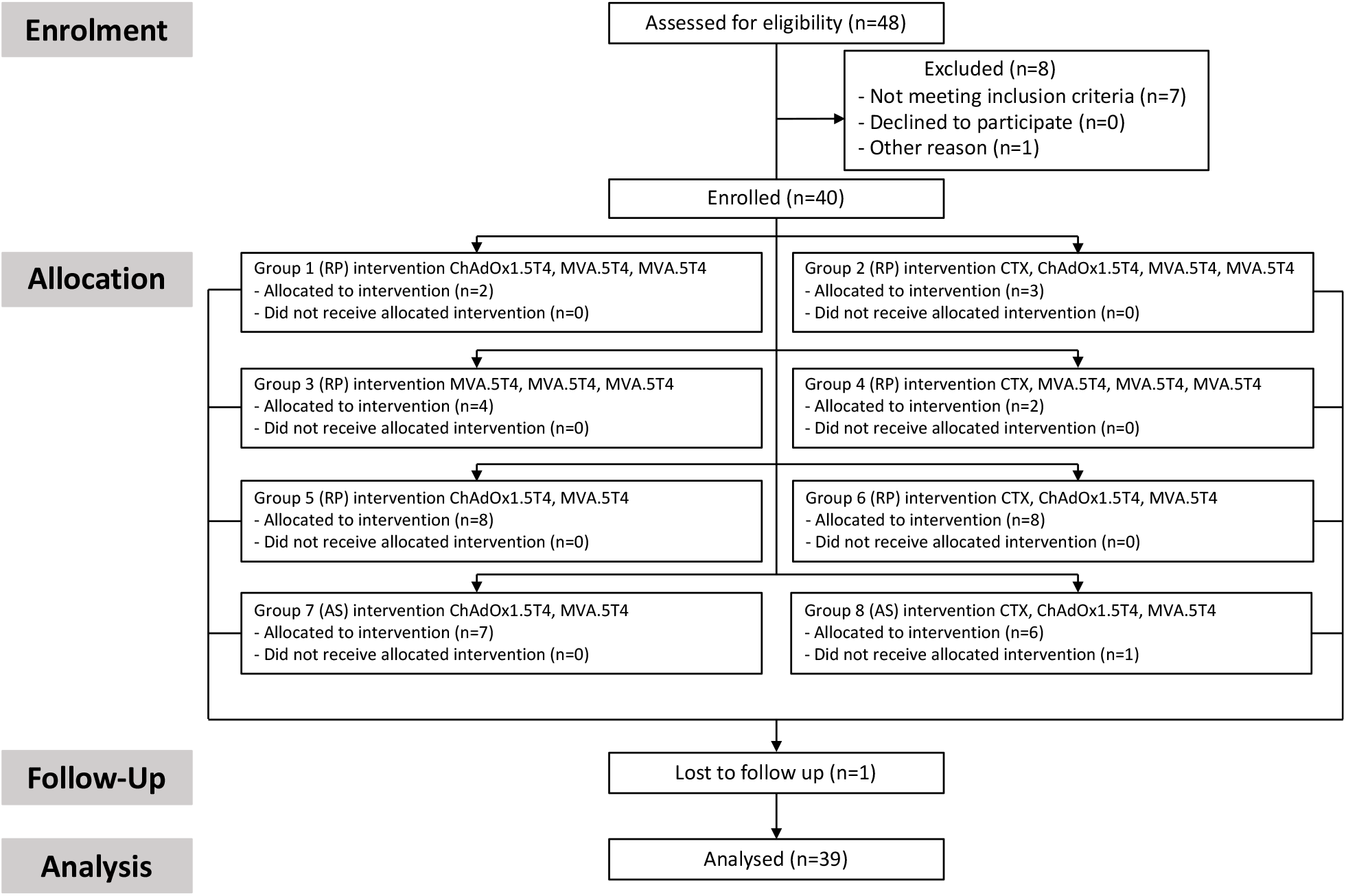
CONSORT flow diagram. Flow chart showing patients participation in VANCE trial study from screening, enrolment, allocation, follow up and analysis. RP: radical prostatectomy; AS: active surveillance; CTX: cyclophosphamide.

ChAdOx1.5T4 and MVA.5T4 vaccines were well tolerated. The majority of reported AEs related to vaccinations were mild in intensity (92%) and were consistent with side effects observed for these vectors in other clinical trials. Mild to moderate pain at injection site was reported by ∼50% of patients. The majority of systemic AEs were graded as mild. Among the common systemic adverse events usually seen post replication-incompetent viral-vectored vaccines, feverishness, myalgia and fatigue were the most frequently reported, affecting 59%, 70% and 72% of individuals respectively, and usually were resolved within 7 days post-vaccination.

There were no reported serious adverse events (SAEs) related to the vaccines. The two reported SAEs, hospital admissions due to an episode of haematemesis and an aortic surgery caused by ascending aortic aneurysm, were considered unrelated to the vaccines.

### Vaccine-induced T cell responses in the blood

25 of 39 (64%) patients who completed the vaccination course and were eligible for analysis mounted 5T4-specific T cell immune responses at one or more blood sampling timepoint, as measured by *ex vivo* IFN-γ enzyme-linked immunospot (ELISpot) assay, with a mean peak response of 198 SFC per million PBMCs (Fig. 2A). The detected *ex vivo* 5T4-specific immune T cell reactivity was attributed to *de novo* induced responses, and only 2 of 39 patients demonstrated pre-existing 5T4 responses, which increased after vaccination (Fig. 2A). The standard heterologous regimen (ChAdOx1-MVA-MVA 5T4, 4-week interval) induced an antigen specific response in 4 of 5 (80%) patients, and the standard homologous regimen (MVA-MVA-MVA 5T4, 4-week interval) in 4 of 6 (66.7%) patients, with an overall immune response rate of 73%. In the accelerated heterologous regimen (ChAdOx1-MVA 5T4, 1-week interval), 17 of 28 (61%) patients showed cellular responses to 5T4 (Fig. 2B). The magnitude of antigen-specific immune response was not significantly different between standard homologous and heterologous vaccination regimens, perhaps due to the small number of patients in these arms. In other indications, the T cell immunogenicity of ChAd-MVA regimens is consistently higher than that of homologous MVA regimens (*16, 17*). Also, comparable frequencies of antigen-specific T cells were observed following the standard heterologous and accelerated heterologous immunisation schedules (Fig. 2B), as has been observed for other vaccinations such as Ebola (*11*). The CTX preconditioned arms did not show any increase in the magnitude of circulating T cell responses compared to the vaccine only arms (Fig. 2C). Importantly, immunisations against the 5T4 protein elicited T cell responses with broad epitope specificities that changed over time (Fig. 2D).

**Fig. 2.**
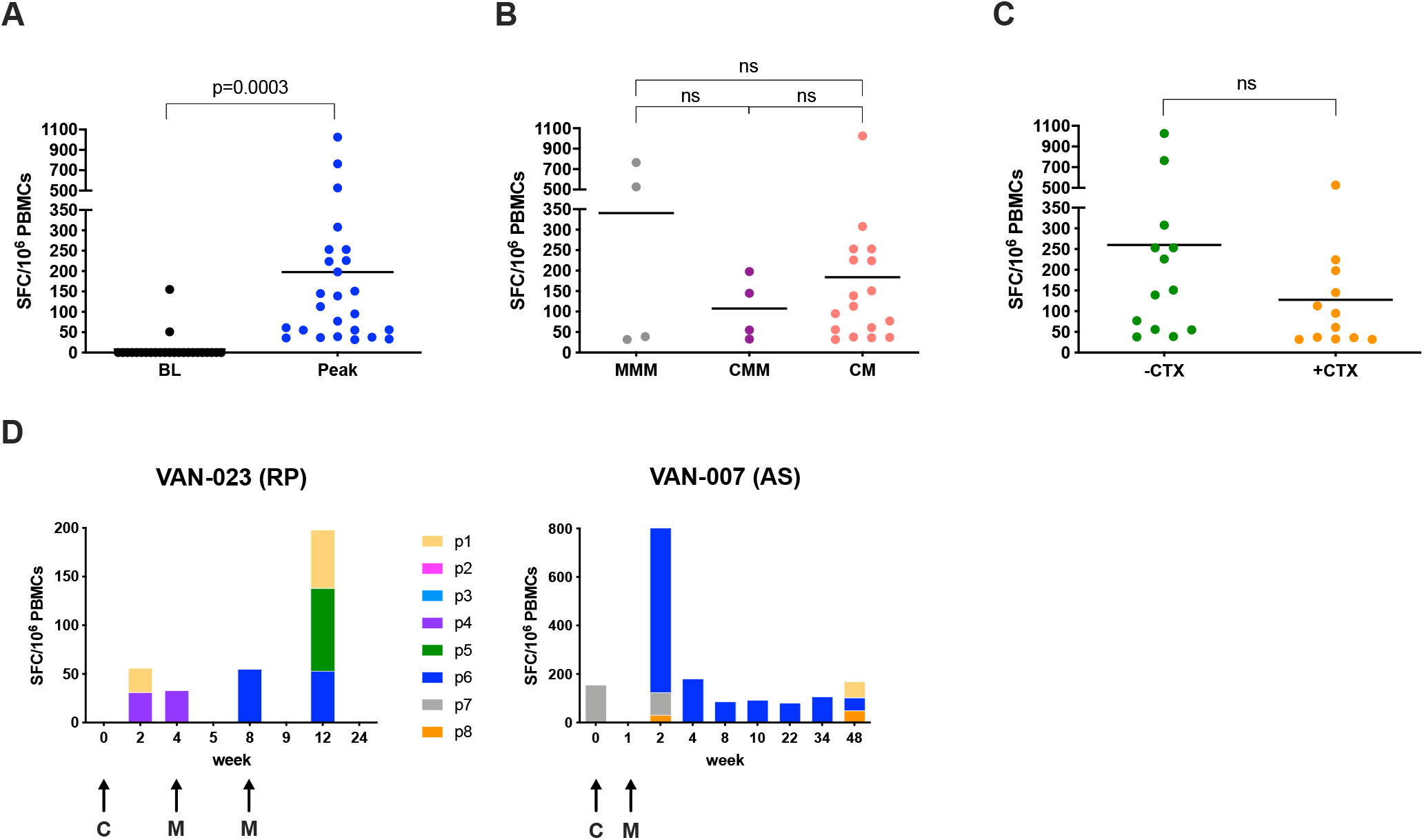
*Ex vivo* IFN-γ ELISPOT responses. Vaccine immunogenicity was assessed by *ex vivo* IFN-γ ELISPOT on freshy isolated PBMCs stimulated with 8 pools of overlapping peptides covering the entire 5T4 protein sequence. Values of spot forming cells (SFC) per million PBMCs correspond to the sum of the responses to the single pools. (**A**) Dot plot representing peak responses for each individual patient compared to baseline (BL), pre-vaccination responses. Lines represent mean; the paired t-test *P* value is shown. (**B**) Comparison of peak responses in patients receiving homologous standard vaccination (MVA-MVA-MVA 5T4: MMM), heterologous standard vaccination (ChAdOx1-MVA-MVA 5T4: CMM) and heterologous accelerated vaccination regimes (ChAdOx1-MVA 5T4: CM), irrespective of CTX preconditioning. Lines represent mean; ns: not significant. (**C**) Comparison of peak responses between patients without CTX preconditioning (-CTX) or receiving CTX course (+CTX) for a week before each immunisation, irrespective of vaccination regimen. Lines represent mean; ns: not significant. (**D**) Time-dependent dynamics of cellular responses to individual 5T4 peptide pools (p1 to p8) in 2 representative patients. Arrows represent time points of vaccination. RP: radical prostatectomy; AS: active surveillance; C: ChAdOx1.5T4; M: MVA.5T4.

26 of 39 (67%) patients also mounted 5T4-specific antibody responses as measured by a relative-quantitative ELISA (*18*). Of note, 5T4 seroconversion occurred in all the patients in the standard vaccination regimen arms compared to the 46% seroconversion rate in patients vaccinated according to the accelerated immunisation schedules (Supplementary Table S2). This finding suggests that a second MVA boost and / or longer intervals between vaccinations are more favourable for induction of antibody responses to 5T4. There was no correlation between the induced 5T4-specific antibody and T cell responses (Supplementary Fig. S2). In order to evaluate T cell immune response kinetics, patients on the same vaccination schedule were pooled irrespective of other variables (i.e. CTX status pre-treatment, and heterologous or homologous vaccination). In patients on the standard schedule, the 5T4-specific T cell response was observed to peaked on two occasions: firstly at 4 weeks after priming, and secondly one week after the second boost (week 9), at a mean of 54 ± 48 SEM and 72 ± 69 SEM SFC per million PBMCs respectively (Fig. 3A). This mirrors the immunogenicity of these vectors in other indications, where responses post-ChAd peak at 2-4 week, and responses post-MVA peak at one week. A peak T cell response in patients assigned to the accelerated regimen was observed one week after the boost at a mean of 80± 27 SEM in the surgical arm, and 131± 85 SEM SFC per million PBMCs in the active surveillance arm (Fig. 3B, C). Following a contraction phase of the immune response, only weak *ex vivo* responses were detectable at later time points, likely because effector cells diminish while less activated central memory cells increase (*10*). Indeed, the repertoire of memory 5T4-specific T cells could be expanded by short-term *in vitro* culture in the presence of the total 5T4 peptide pool from blood samples collected 4-6 months after vaccination. Of note, frequencies of expanded 5T4-specific T cells from blood samples collected prior to vaccination from the same patients were usually much lower (Fig. 3D).

**Fig. 3.**
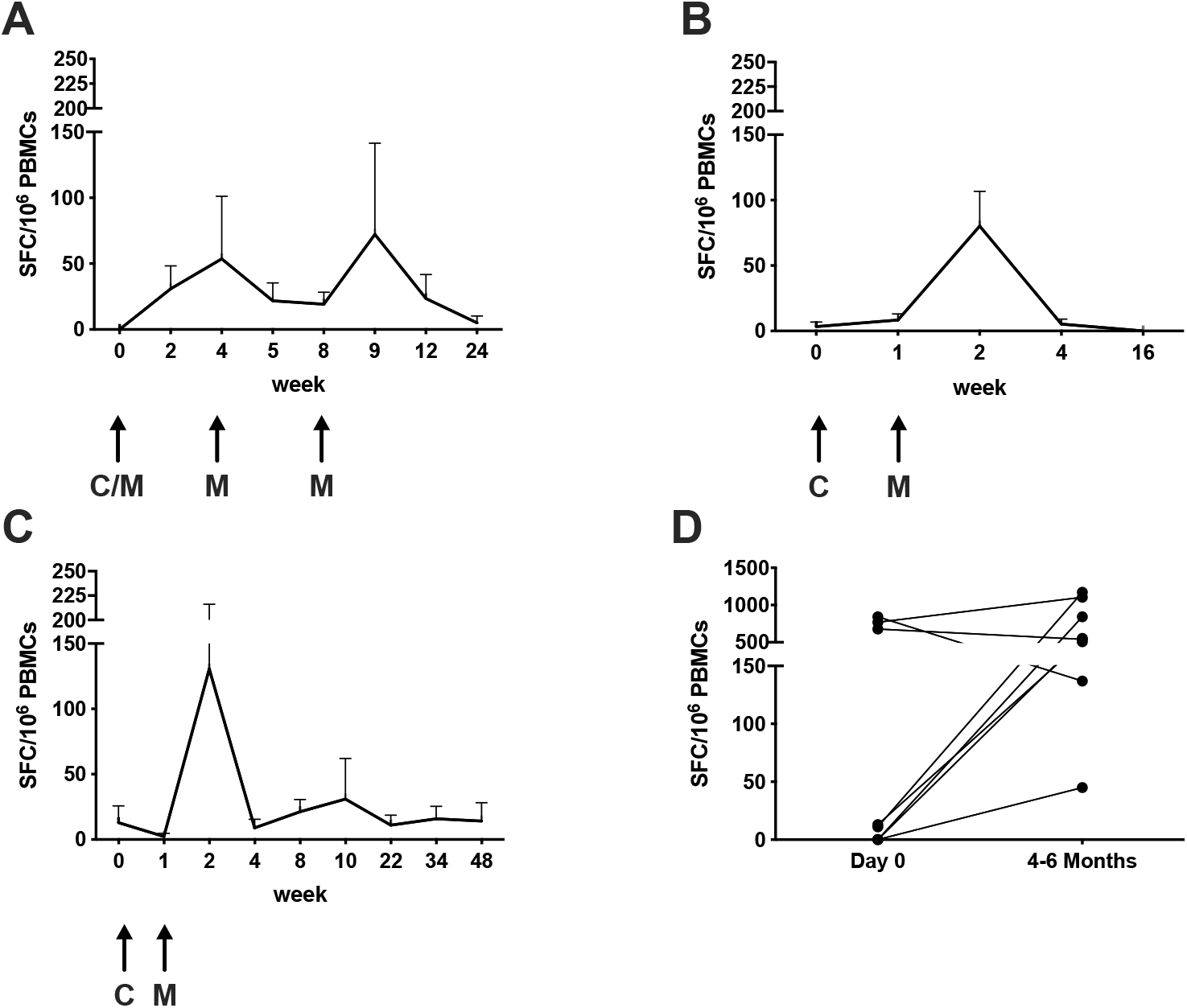
Kinetics of 5T4 specific responses. Antigen-specific responses were measured before vaccination, and at different time points post-vaccination, by *ex vivo* IFN-γ ELISpot assay on freshy isolated PBMCs. Mean (±SEM) immune responses to 5T4 antigen over time are shown irrespective of CTX preconditioning: (**A**) in the standard vaccination regimen arms; (**B**) in surgical patients receiving the accelerated vaccination regimen; and (**C**) in active surveillance patients receiving the accelerated vaccination regimen. Arrows represent time points of vaccination. (C/M: ChAdOx1.5T4 or MVA.5T4; C: ChAdOx1.5T4; M: MVA.5T4). (**D**) Freshy isolated PBMCs from several patients were cultured for 2 weeks in the presence of low dose hIL-2 and the total 5T4 peptide pool. IFN-γ ELISpot assay was performed on cultured cells and baseline responses were compared to responses measured at weeks 16 to 24.

Vaccine-induced T cell responses were further characterized using flow cytometry, which is a less sensitive assay than ELISpot and therefore requires an expansion of the relatively infrequent 5T4-specific T cells. To this end, PBMCs from several patients collected at baseline and at various timepoints during the study have been cultured for 12-14 days in presence of the total 5T4 peptide pool as above. Following *in vitro* stimulation, cell cultures were stained with fluorochrome-labelled antibodies against CD4+ and CD8+ T cell surface markers and intra-cellular IFN-γ and TNF-α cytokines characteristic for effector T cells. The gating strategy is presented in Fig. 4A, and representative plots of IFN-γ+TNF-α+ CD4+ and CD8+ T cells upon 5T4 stimulation are shown in Fig. 4B. 5T4-specific T cell responses in cultured PBMCs were detected in 15 of 21 (71.4%) patients analysed. Among the responders, 5T4-specific CD4+ T cell reactivity was found in 10 (66.7%) patients, CD8+ T cell reactivity in 14 (93.3%) patients and both CD4+ and CD8+ T cell mediated responses in 9 (60%) patients (Fig. 4C). Patients displaying a post-vaccination positive immune response by flow cytometry also had a low percentage of both CD4+ and CD8+ polyfunctional T cells detected at baseline (ranges 0.2-1.8% and 0.01-0.3% respectively). However, these numbers at least doubled after vaccination, reaching as high as 5.3% and 7.8% of 5T4-specific CD4+ and CD8+ T cells at the peak of the response (Fig. 4D). The median fold increase of 5T4-specific T cell responses over baseline values amounted to 4.9 and 4.5 in CD4+ and CD8+ T cells respectively (Fig. 4E). To corroborate the flow cytometry data on induction of both CD4+ and CD8+ 5T4-specific responses after short-term *in vitro* stimulation, PBMCs from selected responders have been retrospectively thawed to perform CD4+ and CD8+ T cell depletions and to test the depleted cell fractions in parallel with the total PBMCs in an *ex vivo* IFN-γ ELISpot assay (Supplementary Materials and Methods). As shown in Supplementary Table S3, 9 patients, including one non-responder, have been analysed to discriminate between CD4+ and CD8+ T cell responses *ex vivo*. Among the responders, only 4 maintained the response after freeze/thawing and we could observe CD4+ only mediated, CD8+ only mediated, and both CD4+ and CD8+ mediated T cell responses, consistent with the results observed by flow cytometry.

**Fig. 4.**
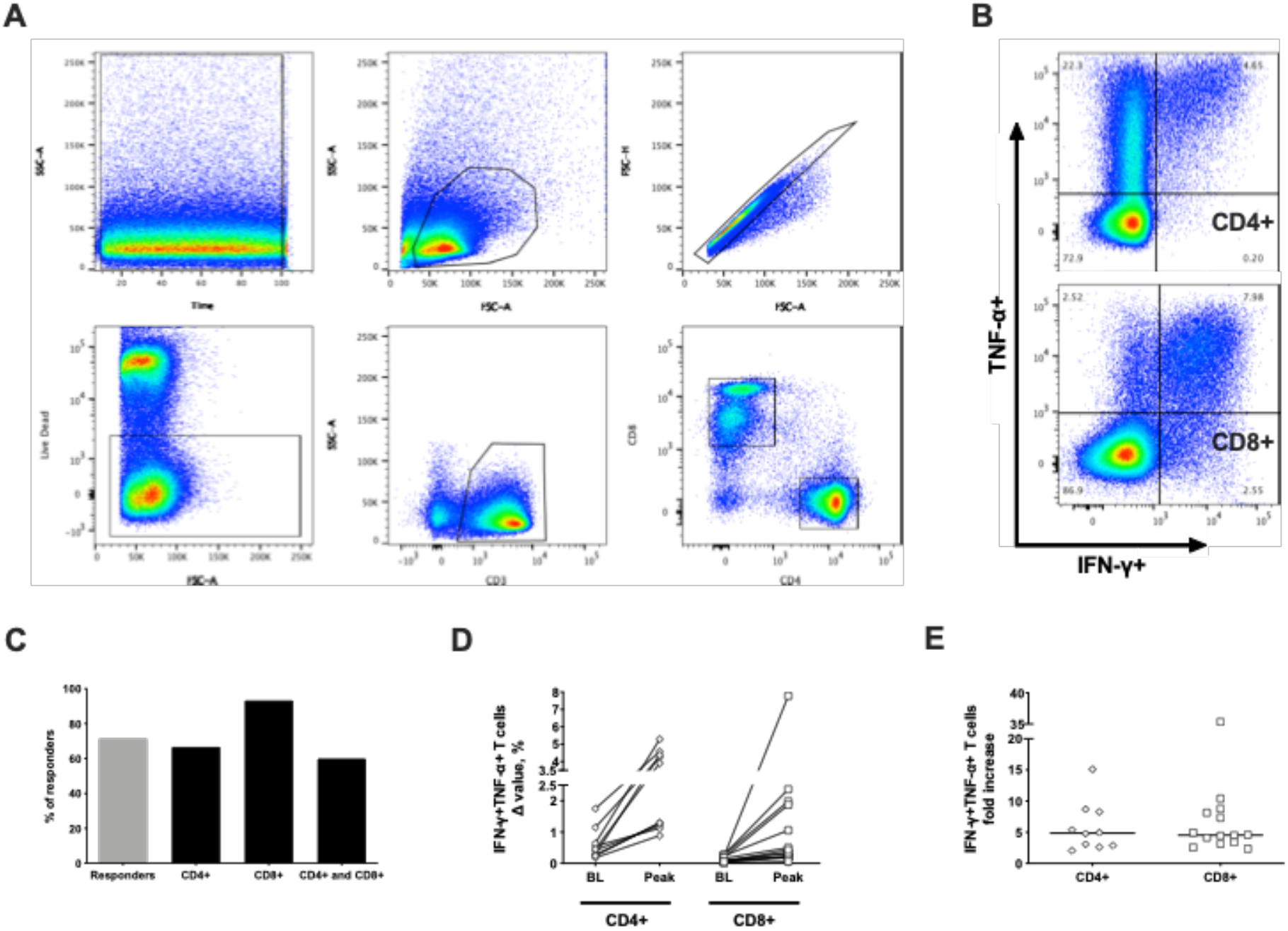
Flow cytometry analysis of 5T4-specific immune responses induced by vaccination. Freshly isolated PBMCs were cultured for 2 weeks in the presence of low dose hIL-2 and the total 5T4 peptide pool to expand relatively infrequent 5T4-specific T cells. Flow cytometric analysis was performed on cultured cells at different time points, and the percentage of polyfunctional CD4+ and CD8+ secreting IFN-γ+ and TNF-α+ was calculated by subtracting background responses (Δ value) and compared to corresponding pre-vaccination values. (**A**) Representative gating strategy. From left to right, top to bottom: Time – Lymphocytes – Single cells – Live cells – CD3+ T cells – CD4+ and CD8+ T cells. (**B**) Representative dot plot with percentages showing CD4+ (top) and CD8+ (bottom) T cells secreting IFN-γ+ and TNF-α+. (**C**) Percentages of evaluated patients showing a positive response in cultured PBMCs (grey bar) and relative percentages of positivity for CD4+, CD8+, and both CD4+ and CD8+ among responders (black bars). (**D**) Baseline Δ value percentages of IFN-γ+ and TNF-α+ CD4+ and CD8+ T cells are compared to corresponding peak Δ value percentages obtained after vaccination in each individual patient analysed. BL: baseline. (**E**) Fold increase of peak Δ value percentages over baseline of polyfunctional CD4+ and CD8+ T cells induced by vaccination are shown. Lines represent median.

### Vaccine-induced T cell responses in the prostate

In order to assess the immune response to the vaccine antigen in the target prostate gland, post-vaccination tissue cores from radical prostatectomy specimens (*19*) and prostate biopsy samples from active surveillance patients were cultured *in vitro* in the presence of the total 5T4 peptide pool and high dose hIL-2, in order to expand the TILs and enrich them for 5T4-specificity prior to flow cytometric analysis. Immune cell expansion rates and CD4+ to CD8+ T cell ratios varied between patients, as well as between tissue fragments derived from the same surgical or biopsy specimen. Representative dot plots of expanded CD4+ and CD8+ T cells are shown in Fig. 5A. The flow cytometric analysis of TILs was similar to that performed on cultured PBMCs. 5T4-specific T cell responses could be detected in the prostate samples of 13 of 17 (76.5% response rate) patients analysed. Overall, prostate tissue analysis demonstrated the presence of 5T4-specific CD4+ T cell reactivity in 8 patients (61.5% of responders), CD8+ T cell reactivity in 11 patients (84.6% of responders) and both CD4+ and CD8+ T cell mediated responses in 6 patients (46.2% of responders) (Fig. 5B). Percentages of TILs secreting both IFN-γ and TNF-α measured in 2 representative patients are shown in Fig. 5C, presenting a comparison between unstimulated and 5T4-stimulated cells.

**Fig. 5.**
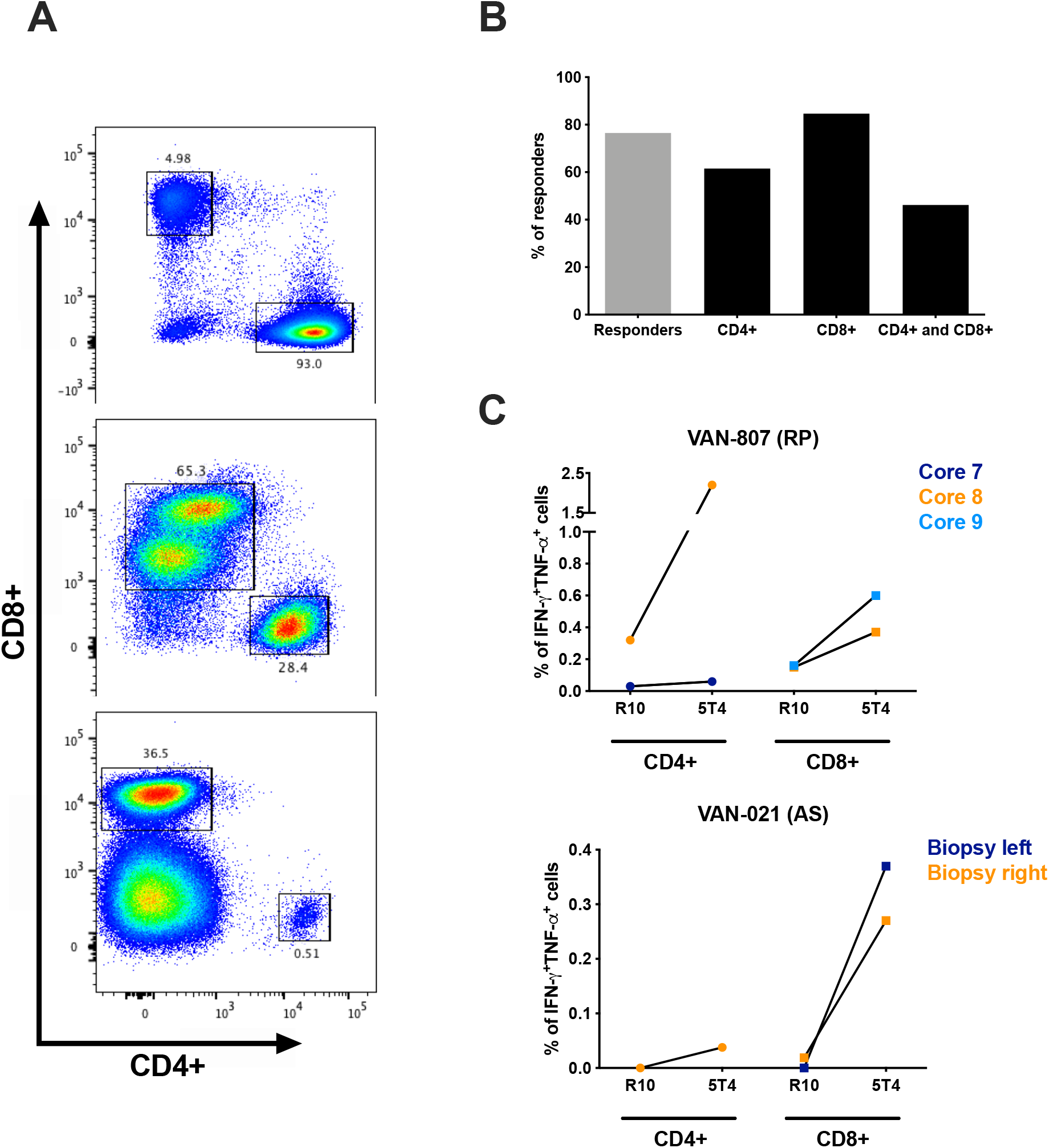
Flow cytometry analysis of 5T4 specific immune responses in the prostate tissue. Post vaccination prostate biopsy samples and radical prostatectomy tissue specimens were cultured in the presence of high dose hIL-2, and were stimulated with the total 5T4 peptide pool to expand 5T4-specific T cells derived from the prostate gland. Flow cytometric analysis was performed on expanded TILs a week after 5T4 stimulation. (**A**) Representative flow dot plot showing percentages of gated CD4+ and CD8+ TILs obtained from expanded cultures in different samples. (**B**) Percentages of evaluated samples showing a positive response in cultured TILs (grey bar) and relative percentages of positivity for the CD4+, CD8+ T cell and both CD4+ and CD8+ T cell subsets (black bars). (**C**) Percentages of IFN-γ+ and TNF-α+ TILs (CD4+ and CD8+) in unstimulated cultures (R10: medium only) are compared to the corresponding 5T4-stimulated cultures (5T4) in 2 representative patients (VAN-807 and VAN-021). RP: radical prostatectomy; AS: active surveillance.

To evaluate post-treatment changes in the immune cell infiltration using immunohistochemistry (IHC), tissue sections from archival diagnostic prostate biopsies and matched post-vaccination on-study biopsies from active surveillance patients were stained with anti-CD3 and anti-CD8 monoclonal antibody, and densities of CD3+ and CD8+ T cells were assessed by digital image analysis as previously described (*20*). An increase in CD3+ and CD8+ T cell density was observed in the post-vaccination prostate gland in some of the analysed patients. In particular, an increase equal or higher than 1.5-fold in CD3+ and CD8+ T cell density post-treatment was detected in 2 out of 11 patients (18%) and in 4 out of 11 patients (36%) respectively (Supplementary Fig. S3).

IHC analysis was also performed on 12 available radical prostatectomy tissue sections from patients in Groups 1-4, and both CD3+ and CD8+ T cell infiltration was detected in resection samples (Supplementary Table S4). However, T cell densities in the prostate gland following vaccinations could not be reliably compared with the pre-treatment archival biopsy samples due to technical issues.

### Serum PSA changes following vaccination

The serum PSA level is commonly used as a surrogate marker of prostate cancer treatment efficacy. Potential treatment effects of the vaccine on prostate cancer were assessed by measuring serum PSA throughout the trial. In the majority of patients in the surgical arms PSA could only be assessed at a few time points prior to radical prostatectomy, as this intervention ordinarily causes the PSA to drop to very low or undetectable levels. However, patients on the active surveillance programme could be followed up for 48 weeks, thus changes in PSA were monitored in these patients for an extended time period. Fig. 6A shows PSA changes over time in the 12 patients in the active surveillance arms. Although there was some disparity in the PSA kinetics between patients, the overall trend showed an unexpected increase in serum PSA concentration after ChAdOx1.5T4 prime with a peak at week 4, with PSA decreasing to pre-vaccination levels by week 22 (Fig. 6B). Of note, median peak levels of PSA after vaccination were significantly higher than median baseline levels (8.25 ng/ml versus 4.16 ng/ml) (Fig. 6C). Analysis of the maximum PSA change detected at any given time point post-vaccination demonstrated that 50% of patients experienced an increase of 17-52% over the baseline. 25% of patients had ≥100% increase in PSA concentration, and 25% of the patients had a PSA drop of 12-38% compared to pre-vaccination values (Fig. 6D).

**Fig. 6.**
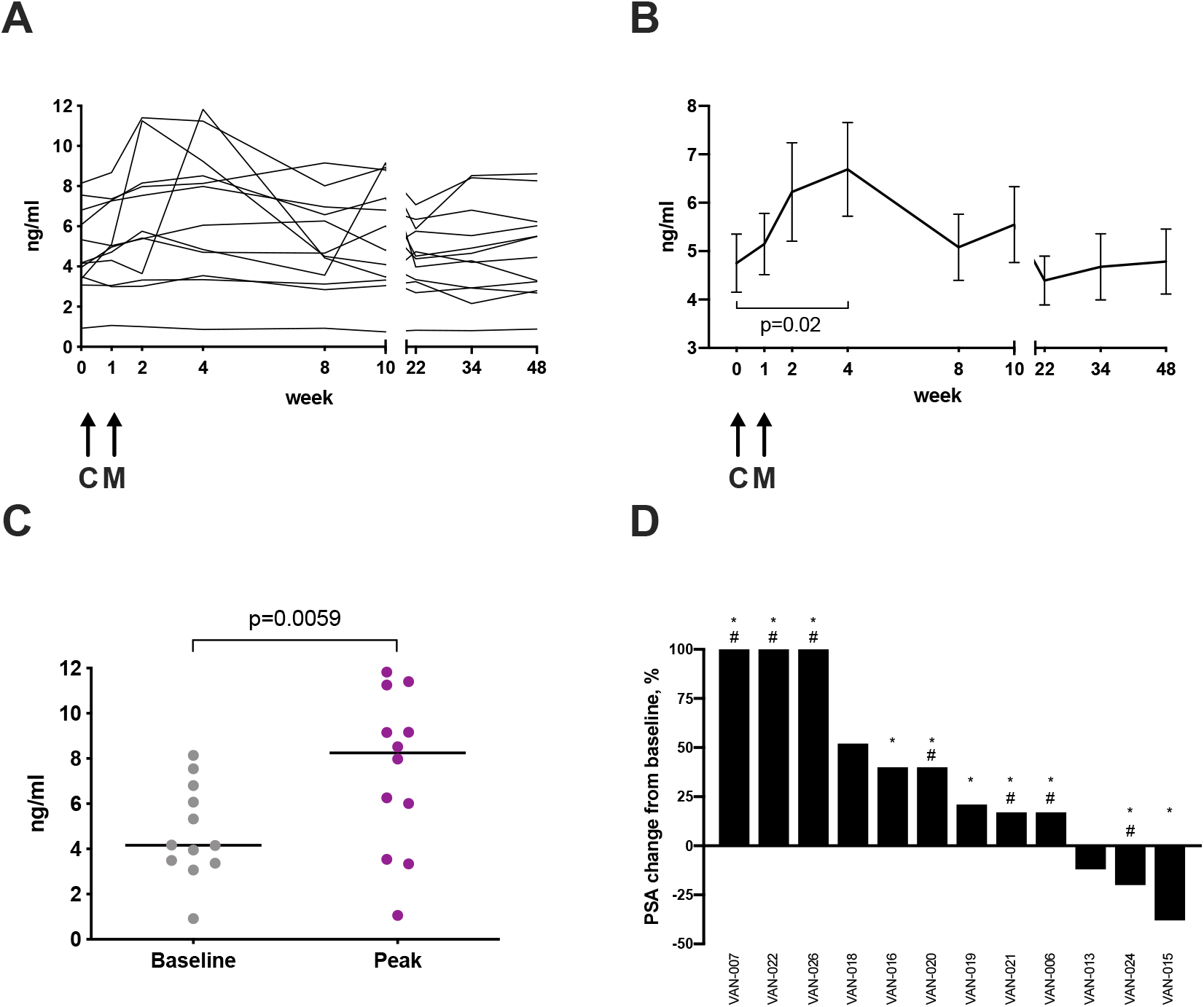
PSA levels in the active surveillance arms. Serum PSA concentration was measured throughout the study. Changes in PSA concentration are shown for each individual patient in the active surveillance arms (**A**), and as mean concentrations±SEM (**B**); Mean PSA levels at each time point; paired t-test value *P* = 0.02. Arrows represent time points of vaccination. C: ChAdOx1.5T4; M: MVA.5T4. (**C**) A scatter dot plot graph representing baseline PSA concentrations compared to corresponding peak concentrations measured post vaccination in active surveillance patients. Lines represent median; paired t-test value *P*=0.006. (**D**) Waterfall plot representing the highest PSA change from baseline, expressed as percentage of baseline value, measured at any given timepoint in patients on the active surveillance arms (patient trial IDs are given). * represents a systemic immune response to the vaccine measured either by *ex vivo* IFN-γ ELISpot on freshly isolated PBMCs or flow cytometry on short-term stimulated PBMCs. # indicates a CD8+ T cell local immune response to the vaccine measured either by flow cytometry on short-term stimulated TIL cultures or by IHC of post vaccination biopsies (CD8+ T cell density ≥1.5-fold pre-vaccination biopsies).

The increase of ≥100% in serum PSA levels observed in 3 patients was associated with both peripheral and local biomarkers of vaccine immunological activity, such as presence of polyfunctional 5T4-specific CD8+ and CD4+ T cell responses in the blood as measured by *ex vivo* ELISpot and flow cytometry of short-term cultured PBMCs, expansion of 5T4-specific polyfunctional T cells from fresh prostate biopsy samples, and a ≥1.5-fold increase in CD8+ T cell infiltration in post-treatment prostate biopsies compared to baseline (Table 1). T cell densities, as measured by IHC, of 2 of these 3 patients are shown in Fig. 7, as well as representative images of IHC staining of the respective prostate biopsies. The TIL and IHC data for the third patient (VAN-022) were not available as his on-study biopsy was not performed for clinical reasons.

**Table 1.**
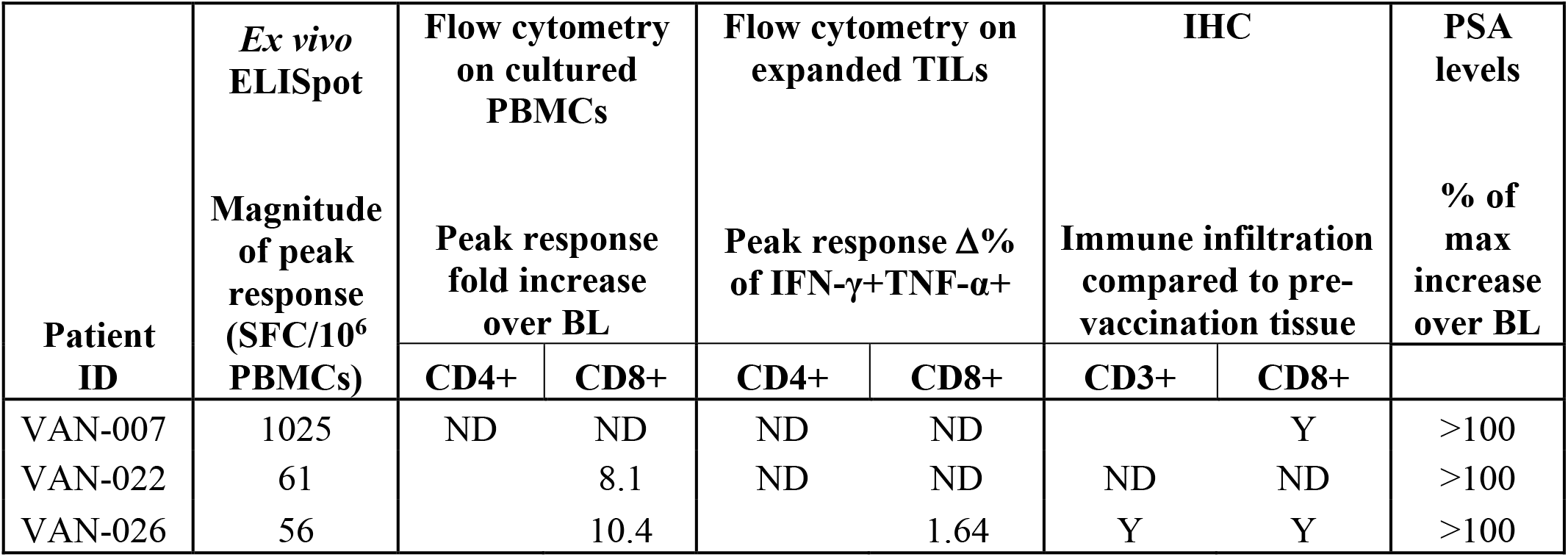
Comparative immune T cell reactivity against the 5T4 antigen as measured by functional assays in the blood and the prostate along with density of T cell infiltration by IHC in 3 selected patients with ≥100% increase in serum PSA. BL – baseline, ND – not done, N - No, Y – Yes.

| Patient ID | *Ex vivo* ELISpot<br>Magnitude of peak response (SFC/$10^6$ PBMCs) | Flow cytometry on cultured PBMCs | | Flow cytometry on expanded TILs | | IHC | | PSA levels |
| --- | --- | --- | --- | --- | --- | --- | --- | --- |
| | | Peak response fold increase over BL | | Peak response $\Delta\%$ of IFN-γ+TNF-α+ | | Immune infiltration compared to pre-vaccination tissue | | % of max increase over BL |
|  |  | CD4+ | CD8+ | CD4+ | CD8+ | CD3+ | CD8+ |  |
| VAN-007 | 1025 | ND | ND | ND | ND |  | Y | >100 |
| VAN-022 | 61 |  | 8.1 | ND | ND | ND | ND | >100 |
| VAN-026 | 56 |  | 10.4 |  | 1.64 | Y | Y | >100 |

**Fig. 7.**
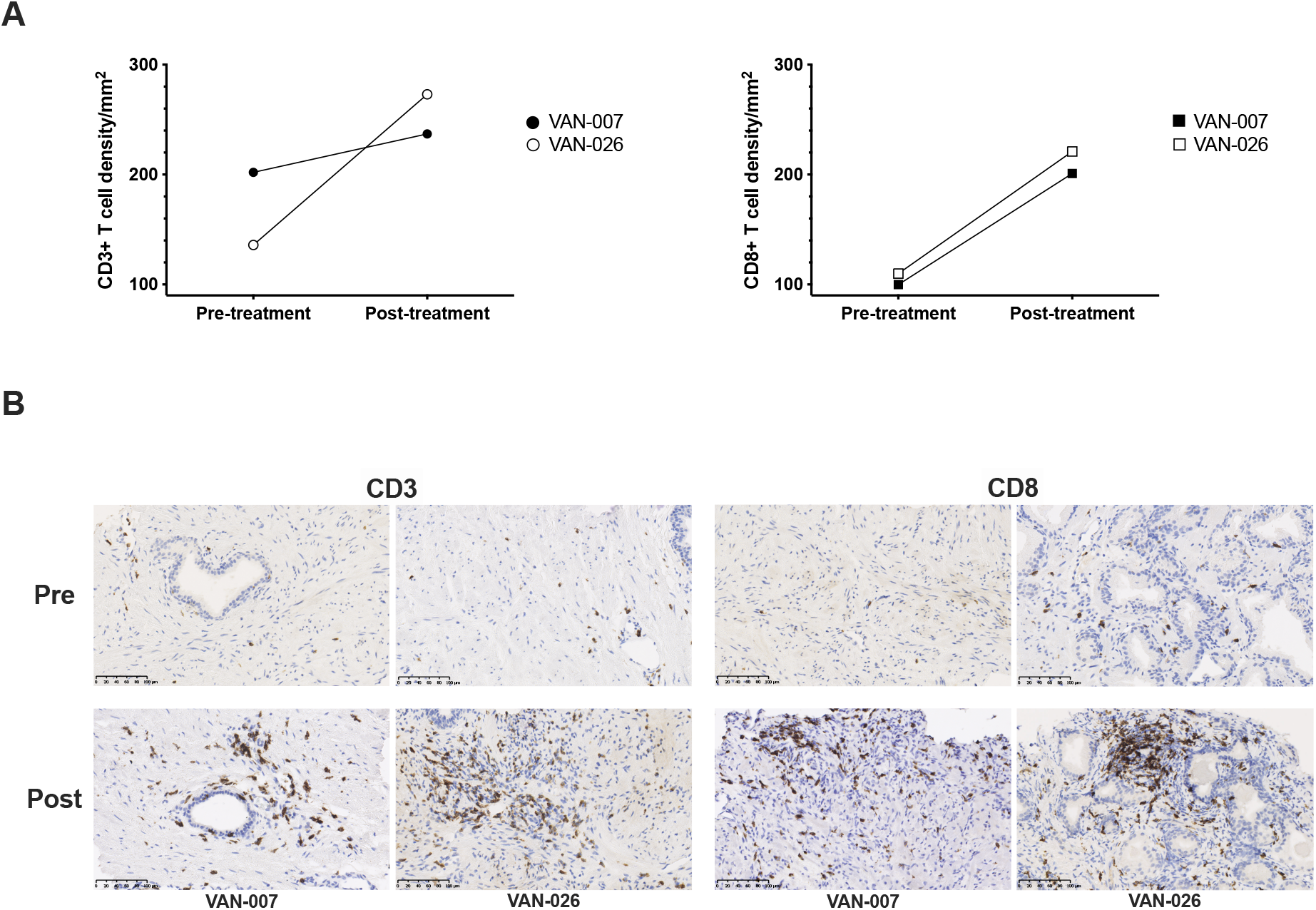
T cell infiltration in the prostate. Archival diagnostic prostate biopsies and matched on-study prostate biopsies taken at week 10 from selected patients in the active surveillance arms were tested for T cell infiltration by immunohistochemistry (patient trial IDs are given). Formalin-fixed paraffin-embedded sections were stained with anti-CD3 and anti-CD8 antibodies and cell densities were calculated. (**A**) Comparison of CD3+ (left) and CD8+ (right) T cell densities, expressed as number of cells per mm^2^, and (**B**) representative pictures of CD3 and CD8 expression between pre-treatment and post-treatment biopsies in 2 patients who had ≥100% increase in serum PSA levels post-vaccination. Brown regions indicate immunoreactivity. Scale bar is shown (100μm).

## Discussion

In this study, the safety profile and immunogenicity of the first-in-man ChAdOx1-MVA heterologous prime-boost vaccination against the 5T4 tumour antigen, in low- and intermediate-risk prostate cancer patients, has been evaluated. The rationale for selecting this patient cohort is that immunotherapy at early stages of the disease should result in greater clinical benefit, as anti-tumour immune responses need time to develop, and the tumour burden may be comparatively low, and more accessible to cytotoxic T cells, which are the major players in anti-tumour protection. An indication of improved benefit in early vaccine therapy in prostate cancer patients comes from observations in the IMPACT trial, where efficacy of Sipuleucel-T treatment was particularly pronounced in patients with lower PSA baseline values (*21*). Another important consideration was the availability of post-vaccination fresh prostate tissue from biopsy and/or surgical resection specimens, which enabled an assessment of the effect of vaccination on the target organ in addition to the evaluation of induced immune responses in the periphery.

The heterologous ChAdOx1-MVA 5T4 vaccine was well tolerated in both the standard and accelerated regimens. The well-tolerated safety profile was in agreement with previous reports for these viral vectors (*8, 22, 23*), with only two serious AEs reported over the duration of the study, both of which were unrelated to the investigational products.

Vaccines induced *de novo* 5T4-specific immunity in each of the eight arms of the study, and significantly boosted two pre-existing responses. Overall, 64% of patients were found to have responded to vaccination on at least one time point during the study, with some responses detected as early as after a single priming immunisation. To our knowledge, this is the best vaccine-induced immune response rate obtained to date in any clinical trial in prostate cancer patients measured by *ex vivo* IFN-γ ELISpot assay.

Breaking tolerance to self-antigens by cancer vaccines has been a challenge that has hampered progress in the cancer immunotherapy field for years, and available information on induced T cell responses in prostate cancer trials remains sparse. The response rate achieved in the VANCE trial, together with a magnitude of response ranging between 30 and 1025 SFC per million PBMCs, can be considered noteworthy when compared to immunotherapy responses measured in other clinical trials on prostate cancer patients.

The closest comparator for the ChAdOx1-MVA 5T4 vaccine is TroVax, a homologous MVA.5T4 vaccination regimen in the prostate cancer setting (*22*). In the reported phase II trial, 5T4-specific T cell responses were induced in 38% of hormone refractory prostate cancer patients over the course of 11 TroVax immunisations. ProstVac treatment, the poxvirus-based vaccine targeting PSA, induced PSA-specific T cell responses in 57% of patients across six clinical trials. But the magnitude of the response measured 4 weeks after the last immunisation was in the range of 10 to 203 SFU per million PBMCs, with a median response of 30 SFU per million PBMCs. The low T cell responses to the vaccine antigen was interpreted as a reflection of sub-optimal timing for blood sampling and/or trafficking of PSA-specific T cells to the tumour site (*4*). Of note, the highest frequency of responses across 6 ProstVac trials was detected in localized prostate cancer patients. These data suggest that a smaller tumour burden, as well as a generally better immunological fitness of the patients, might positively influence the response rate to cancer vaccines, as observed in the present VANCE study.

The licenced cell-based immunotherapy Sipuleucel-T induced T cell responses to the target antigen, a recombinant fusion protein of PAP and GM-CSF (PA2024), in 48% of treated mCRPC patients compared to 6% in the control group. However, the response rate to the PAP antigen itself was in the range of 6-8% in both treated and control patients (*3*), which suggests that much of T cell reactivity against the recombinant protein may have been directed to the junctional foreign antigens. In another clinical trial, a DNA vaccine encoding PAP and GM-CSF (pTVG-HP) was assessed for immunogenicity in prostate cancer patients with biochemical recurrence. This vaccine only yielded a low response rate of 14% (*24*). A follow-up study demonstrated that multiple immunisations were necessary for robust responses, as the majority of patients showed detectable PAP-specific T cells after a course of 6 vaccinations (*25*). The pTVG-HP vaccine has been evaluated more recently in mCRPC in combination with anti-PD1 (pembrolizumab) (*26*). The reported T cell responses were likely to be predominantly CD4+ T cell mediated, as the PAP protein induced much high frequencies of PAP-specific T cells by IFN-γ ELISpot compared to the PAP peptide antigen. Also, the responses to PAP peptides were detected after 48-72 hours *in vitro* re-stimulation, in contrast to the standard 18-hour assay used here. Several other prostate cancer vaccine candidates are currently being tested in clinical trials in combination with PD-1 checkpoint inhibitors (NCT02325557, NCT02616185).

RNA-based vaccination approaches have also been evaluated in mCRPC (*27*). The self-adjuvanted mRNA vaccine CV1903 against 4 prostate antigens induced T cell responses in 55% of patients. These responses were detected after 6 doses of the vaccine and reported as a cumulative response rate measured by MHC-class I tetramer staining, ICS and IFN-γ ELISpot. A comparable magnitude of 5T4-specific response was observed across all vaccination regimens in the VANCE study, with no detectable effect from CTX preconditioning. This observation was unexpected, as low dose CTX may selectively suppress regulatory T cell (Treg) number and functionality, without influencing other T cell subsets, and this effect would have the potential to uncover high-avidity tumour-specific T cells (*28*). However, our observation of little impact from CTX is corroborated by other studies. For example, depletion of Tregs with low-dose cyclophosphamide prior to vaccination did not increase immune responses generated by MVA.5T4 vaccination in metastatic colorectal cancer patients (*29*). The hypothesis was that CTX might independently improve the efficacy of vaccination without influencing the immunogenicity of the vaccines. In the majority of VANCE patients, the induced *ex vivo* T cell responses measured by an IFN-γ ELISpot assay have not been maintained over time. However, the 5T4-reactive memory cells could be expanded *in vitro* 5 months after the last immunisation.

We observed the presence of vaccine-induced systemic CD8+ or/and CD4+ T cell immune responses in both *ex vivo* tests and *in vitro* cultured assays. However, vaccine-driven activation of T cells in the circulation does not always predict local responses within the target organ. Traditionally, post-vaccination immune responses in the prostate gland have been investigated exclusively using IHC. As an example, staining of radical prostatectomy specimens after vaccination with Sipuleucel-T showed an accumulation of CD4+ and CD8+ T cells at the tumour interface (*30*). Farsaci *et al*. have also showed an increase in CD4+ and CD8+ T cells within the tumour parenchyma in all patients receiving ProstVac vaccination (*31*). However, an influx of immune cells into the prostate gland may also be caused by other interventions. For example, CD4+, and to a lesser extent CD8+, T cell immune infiltration into the prostate gland has been observed after androgen ablative therapy (*32*). Interestingly, in contrast to observations seen in other malignancies, a positive correlation between the density of tumour infiltrating CD8+ T cells and improved clinical outcome remains controversial in prostate cancer (*33*). On one hand, it has been suggested that in comparison to absent or decreased immune infiltrates, abundant TILs in the prostate gland is associated with a higher survival probability (*34*). On the other hand high numbers of TILs within the tumour have been demonstrated to be predictive of an increased risk of tumour recurrence in patients undergoing radical prostatectomy, with an expansion of the CD4+ T cell infiltrate being associated with a poor outcome (*35, 36*). In addition, a strong TIL presence was an independent predictor of short PSA recurrence-free survival (*37*). More recently, higher levels of primarily CD4+ T cells and, to a lesser extent, CD8+ T cells, appeared to be associated with a worse distant metastasis-free survival (*38*). It may therefore be the case that in addition to enumerating T cells in the prostate tumour, the functional status and tumour specificity of TILs after the vaccination are important characteristics to analyse. To this end, in parallel with the IHC assessment of formalin-fixed paraffin-embedded prostate tissue sections, in the VANCE study we obtained high-quality fresh post-vaccination prostate tissue, in order to study the presence of functional vaccine-specific T cell responses. We could quantitatively assess 5T4-specific polyfunctional TILs (i.e. secreting IFN-γ and TNF-α) from the majority of analysed samples. Of note, 11 of 13 (85%) patients demonstrated presence of functional CD8+ T cells, with over 50% of these patients exhibiting a concomitant CD4+ T cell response. A limitation of our approach is that the TILs subjected to this functional analysis have been cultured *in vitro*, isolated from the natural tumour immunosuppressive microenvironment, and so they might have expanded in a more favourable environment. Another drawback of our analysis is that it was not possible to compare the immune cell phenotype and function in the prostate gland pre- and post-vaccination, due to the unavailability of fresh tissue before study treatment. Nevertheless, and despite the uncertain role of TILs within the prostate gland, we could observe functional vaccine-specific cytotoxic and T helper cell responses within the prostate gland after vaccination, as well as T cell infiltration measured by IHC, which might effectively control prostate cancer development, especially if given in prophylactic vaccine settings.

The presence of 5T4-reactive TILs might also explain the transient increase in PSA levels observed in active surveillance patients post ChAdOx1.5T4 prime and MVA.5T4 boost immunisations. This effect might be the result of a vaccine-induced inflammatory response, and possibly reflects prostate epithelial cell death, following trafficking of 5T4-reactive T cells to the target organ. Although the observed PSA trends post-vaccination were variable, the analysed active surveillance patients experienced a significant rise in PSA level, with a peak observed at week 4 (two weeks after the MVA.5T4 boost). Although the PSA in almost all patients had normalised to pre-vaccination values by the end of the study, the transient PSA increase significantly exceeded the physiological variations of PSA in several patients.

Interestingly, a PSA flare phenomenon followed by dramatic clinical and biochemical responses has been observed after treatment with radium-233 in an extensively pre-treated metastatic prostate cancer (*39*). Similarly, mCRPC patients treated with chemotherapy experienced an initial increase in serum PSA, followed by a decrease of at least 50% when compared to baseline (*40*). PSA dynamics have also been analysed after brachytherapy, and mechanisms behind the observed “PSA bounce” have been evaluated (*41*). The “PSA bounce” measured in the brachytherapy study could not be explained by the effect of radiation alone as it invariably occurred after the isotope had decayed. Therefore, the phenomenon was attributed to the differential immune response to the tumour coupled with the therapeutic effect of radiation, as suggested by the strong T cell infiltration detected within the tumours by IHC analysis of the tissue after therapy. Patients who experienced a PSA bounce had higher densities of CD3+ and CD8+ T cells within the tumour, and this might have contributed to both the PSA increase and the improved clinical outcomes observed. These findings are consistent with our hypothesis that the temporary rise in serum PSA observed in VANCE patients may have been caused by the local T cell reactivity induced by vaccination. In fact, the finding of a PSA bounce can be part of so-called tumour pseudoprogression following immunotherapeutic interventions (*42*).

In conclusion, the heterologous ChAdOx1-MVA 5T4 vaccination regimen in early stage prostate cancer was found to be both safe and immunogenic, with the majority of prostate cancer patients enrolled in this trial mounting *ex vivo* T cell responses to the vaccine-encoded tumour-associated antigen 5T4. Immune responses mediated by polyfunctional CD8+ and CD4+ T cells could be detected following vaccination *ex vivo* in the blood, and following *in vitro* culture of cells obtained from prostate tissue. The transient rise in serum PSA observed post vaccination, which significantly exceeded physiological variation in some patients, supports a possible vaccination effect within the prostate gland.

It remains to be seen if the vaccination strategy presented herein can be effective in later-stage prostate cancer therapeutic settings, and whether it can be effective as a monotherapy in advanced disease, or as part of multi-modality prostate cancer therapy. To address these questions we have started the phase I/II trial ADVANCE, which is currently recruiting patients with intermediate-risk prostate cancer, and patients with advanced mCRPC, to receive ChAdOx1-MVA 5T4 vaccine in combination with nivolumab (NCT03815942).

## Materials and Methods

### Study design

VANCE was a phase I randomised open label study designed to assess the safety and immunogenicity of ChAdOx1-MVA 5T4 vaccine with and without low dose cyclophosphamide in low- and intermediate-risk localised prostate cancer. Participants were enrolled at the Oxford University Hospitals NHS Foundation Trust, and the Sheffield Teaching Hospitals NHS Foundation Trust, in the United Kingdom (UK). Written informed consent was obtained from all participating patients, and the trial was conducted in accordance with the principles of the Declaration of Helsinki and Good Clinical Practice. This study was approved within the UK by the Medicines and Healthcare Products Regulatory Agency and the South Centre Berkshire Research Ethics Committee.

The trial was registered with the U.S. National Institutes of Health (NIH) Clinical Trials Registry (ClinicalTrials.gov identifier NCT02390063).

The sample size was chosen to allow an initial descriptive report of the safety and immunogenicity of the vaccine in a first-in-human trial.

Randomization was performed by an independent statistician at the Centre for Statistics in Medicine, Oxford University. Allocation was performed using a balanced-block stratified randomization.

### Study population

The target population for this study was men newly diagnosed with low- or intermediate-risk prostate cancer who had chosen radical prostatectomy as their treatment option, and men with low-volume low-risk prostate cancer on an active surveillance programme (AS) and stable for at least a year. Patients eligible for the study had histologically confirmed prostate cancer, PSA ≤ 20 ng/ml, Gleason score ≤ 7, clinical stage disease ≤ T2c, and no evidence of metastases. Exclusion criteria included history of immunodeficiency disease, systemic immunosuppression, allergic response to previous vaccinia virus vaccinations, seropositivity for hepatitis B surface antigen (HBsAg), hepatitis C virus (HCV) or HIV.

### Study products

Design and construction of ChAdOx1.5T4 vaccine has been described previously (*15*). ChAdOx1.5T4 was manufactured to clinical good manufacturing practice (cGMP) by the Clinical Biomanufacturing Facility (University of Oxford, Oxford, UK) in the human embryonic kidney 293 cell line. The vectored vaccine was purified by cesium chloride isopycnic centrifugation and sterile filtered to generate a clinical lot at a concentration of 2.02?×?1011 vp per ml. The ChAdOx1.5T4 vaccine has been administered at the dose of 2.5 × 10^10^ vp. MVA.5T4 vaccine was manufactured and supplied for the trial by Oxford BioMedica Ltd (TroVax®), and has been described previously (*43*). The MVA.5T4 vaccine has been administered at the dose of 2.0 × 10^9^ TCID50 (equivalent to 1.2 × 10^8^ pfu). Both vaccines were administered by intramuscular injection in the thigh. Cyclophosphamide (CTX) (manufactured by Baxter) was self-administered orally at 50mg twice a day for a seven-day cycle before each vaccination.

### Study procedures

Following screening, patients in the surgical arms were randomly allocated to 6 groups. Participants in Groups 1 and 2 received one ChAdOx1.5T4 immunisation followed by two MVA.5T4 boosts 4 weeks apart and underwent radical prostatectomy at week 12 post enrolment. Participants in Groups 3 and 4 received three MVA.5T4 immunisations 4 weeks apart and also underwent radical prostatectomy at week 12 post enrolment. This immunisation schedule was termed a “standard” vaccination regimen. Participants in Groups 5 and 6 were administered with ChAdOx1.5T4 vaccine followed by MVA.5T4 vaccination one week later and underwent radical prostatectomy at week 4 post enrolment. This immunisation schedule was termed an “accelerated” vaccination regimen. Patients on the active surveillance programme were randomised to Groups 7 and 8 and received an accelerated vaccination regimen followed by on-study prostate biopsy at week 10 post enrolment. The participants in Groups 2, 4, 5 and 6 received an oral course of low dose CTX for 7 days prior to each immunisation. The study treatment schedule is outlined in Supplementary Figure S1.

Blood samples were drawn and clinical assessments conducted for safety, immunology and PSA monitoring prior to vaccination at day 0 and at each scheduled clinical time point following enrolment. Solicited local site reactions and systemic symptoms were recorded by participants using paper diaries during the 7-day follow-up period. Unsolicited AEs and SAEs were recorded for 28 days. The severity of AEs was graded using the following criteria: a) mild (short-lived or mild symptoms with no limitation to usual activity); b) moderate (mild to moderate limitation in usual activity); and c) severe (considerable limitation in activity, medication or medical attention required).

Fresh prostate tissue specimens were collected from surgical radical prostatectomy specimens, and from on-study biopsies in active surveillance patients, for analysis of post-vaccination immune cell subsets within the prostate tissue following treatment.

### Endpoints

Primary objectives of the study were safety of ChAdOx1-MVA 5T4 vaccination and the induction of immune responses to the vaccine encoded tumour-associated antigen 5T4 in the blood following treatment. Secondary objectives of the study included comparative immunogenicity assessment of the standard and accelerated vaccination regimens, the effect of CTX pre-conditioning on the magnitude of immune responses, the effect of vaccination on the immune cells in the tumour, and serum PSA change during the study.

### Ex vivo IFN-γ ELISpot

*Ex vivo* IFN-γ ELISpot assay was performed at each time point in order to assess vaccine-induced immune responses. Briefly, 2.5 × 10^5^ freshly isolated PBMCs were plated in 96-well PVDF plates pre-coated with anti-IFN-γ antibody, and stimulated with 8 individual pools each consisting of 10-12 15mer peptides overlapping by 10 amino acids, spanning the complete 5T4 protein sequence. Unstimulated cells were used as a background control and Staphylococcus enterotoxin B (SEB) and CMV/Epstein-Barr/Flu (CEF) peptide mix (Mabtech AB) were used as positive controls. After 18-24 hours incubation, plates were developed and the number of IFN-γ producing cells (SFCs) was evaluated using an automated ELISpot reader (AID). 5T4-specific response was deemed to be positive at any time point when the mean number of SFCs per one million PBMCs was at least 40, and the number of spots per well at least two times the background. If a pre-existing T cell response was detected, a positive response due to vaccination was reported if the post immunisation response was ≥2-fold the response determined before immunization. Positive responses to individual pools were summed to give final SFCs values per million PBMCs.

### In vitro culture of PBMCs and TILs

PBMCs and TILs were cultured *in vitro* to expand 5T4-specific T cells. Briefly, 3 × 10^6^ freshly isolated PBMCs/ml were plated in complete medium with or without total 5T4 peptide pool (5µg /ml) in the presence of recombinant human IL-7 (rhIL-7) (25ng/ml). Every 3-4 days the medium was replaced and rhIL-2 (100U/ml) added to the cultures. PBMCs were harvested after 2 weeks and incubated overnight at 37 °C before further analysis. To obtain TIL cultures, radical prostatectomy specimen tissue cores and prostate biopsies were cut into small pieces and cultured in the presence of rhIL-2 (6000U/ml) as described previously (*44*). TIL medium was replaced every 2-3 days and the total 5T4 peptide pool was added when immune cell cultures entered an exponential growth phase. TILs were typically harvested 1-week post 5T4 stimulation for further analysis.

### Flow cytometric analysis

*In vitro* cultured PBMCs and TILs were incubated with 1µg/ml of co-stimulatory antibodies αCD28/αCD49d (eBioscience) followed by stimulation in 96 well plates with complete medium, SEB (1µg/ml) or total 5T4 peptide pool (5µg/ml). Following a 2-hour incubation, brefeldin A and monensin (BD Bioscience) was added and cells were incubated overnight at 37 °C. PBMCs were stained with surface antibodies, fixed and permeabilized, and incubated with intracellular antibodies (Supplementary Table S5). Dead cells were discriminated by live/dead fixable staining (Life technologies). Samples were acquired on an LSRII flow cytometer and analyzed with FlowJo software.

Patients were deemed to be “responders” when the percentages of IFN-γ+TNF-α+ CD4+ and CD8+ T cells after stimulation were higher than 0.02 and were at least 2-fold the background upon analysis in at least one time point post-vaccination. In addition, for cultured PBMCs the delta values (5T4 value – background value) had to be equal/greater than 2-fold the corresponding baseline (BL) delta value.

### Tissue biopsy evaluation

Archival diagnostic biopsies from active surveillance patients collected within one year prior to enrolment, and on-study template or targeted biopsies obtained at week 10 post enrolment, were evaluated. Tissue sections were stained immunohistochemically for expression of CD3 (clone LN10, Leica) and CD8 (clone C8/144B, Dako) at the ISO 15189 accredited NHS Cellular Pathology Laboratory at Oxford University Hospitals NHS Foundation Trust (Oxford, UK). CD3+ and CD8+ T cells were enumerated according to the Tissue Phenomics approach employing a visual context random forest algorithm (45) by Definiens GmbH (Munich, Germany). Epithelial and stromal regions were segmented via an in-house, pre-trained convolutional neural network (46), and cell densities were calculated as the number of marker positive cells per reference area (mm2).

### Data presentation and statistical analysis

Safety data is presented according to the frequency, severity and duration of solicited local and systemic reactogenicity signs and symptoms for seven days following vaccination. Unsolicited AEs and SAEs were recorded for four weeks post vaccination. Statistical analysis of immunogenicity data was conducted using GraphPad Prism version 8 (GraphPad Software Inc., San Diego, CA, USA). Comparisons between datasets were performed using non-parametric tests, as detailed in the figure legends. All *P* values are two-sided and considered statistically significant if *P* < 0.05.

## Data Availability

The data that support the findings of this study are available from the corresponding author, IR, upon reasonable request.

## Supplementary Materials

### Materials and methods

Fig. S1. VANCE study design.

Fig. S2. Correlation analysis of vaccination induced T cell and antibody responses.

Fig. S3. T cell infiltration in the prostate gland of vaccinated active surveillance patients

Table S1. Baseline demographics and disease characteristics of VANCE study.

Table S2. 5T4- and MVA-specific antibody responses in plasma of vaccinated patients.

Table S3. *Ex vivo* IFN-γ ELISpot results on frozen PBMCs depleted of CD4+ or CD8+ T cells.

Table S4. T cell density in the prostate gland of vaccinated surgical patients measured by IHC.

Table S5. Antibody panel for flow cytometry staining of stimulated cultured PBMCs and TILs.

## Acknowledgments

We thank the trial participants and their families for their involvement in this study, the study teams at the local sites for assistance in the execution of this clinical trial, the Oxford Centre for Histopathology Research for processing samples for IHC. We acknowledge support of Clare Verrill by the NIHR Oxford Biomedical Research Centre. We are grateful to Oxford Biomedica Ltd for supplying the MVA.5T4 vaccine.

## Author contributions

Trial conception and design: FH, AVSH, IR

Experimental design, data analysis and interpretation: FC, AH, IR

Laboratory data generation: FC, EP, AM, GS, RH, SV

Provision of patients: RB, LC, JH, JC, FH

Collection of biological samples or clinical data: CV, IP, MB, CM, AP, RM, SK, SM

Protocol development: RB, LC, PR, TE, FH, AVSH, IR

Administrative support: AB Manuscript writing: FC, IR

Manuscript review: RB, CV, AM, GS, RH, SV, PR, TE, JC, FH, AVSH

Final approval of manuscript: All authors

## References

1. M. B. Culp, I. Soerjomataram, J. A. Efstathiou, F. Bray, A. Jemal, Recent Global Patterns in Prostate Cancer Incidence and Mortality Rates. European urology 77, 38–52 (2020).

2. H. C. Kantoff PW, Shore ND, Berger ER, Small EJ, Penson DF, Redfern CH, Ferrari AC, Dreicer R, Sims RB, Xu Y, Frohlich MW, Schellhammer PF, IMPACT Study Investigators. Sipuleucel-T immunotherapy for castration-resistant prostate cancer. N Engl J Med. 363, 411–422. (2010).

3. N. A. Sheikh et al., Sipuleucel-T immune parameters correlate with survival: an analysis of the randomized phase 3 clinical trials in men with castration-resistant prostate cancer. Cancer immunology, immunotherapy : CII 62, 137–147 (2013).

4. J. L. Gulley et al., Immune Impact Induced by PROSTVAC (PSA-TRICOM), a Therapeutic Vaccine for Prostate Cancer. Cancer Immunology Research 2, 133141 (2013).

5. J. L. Gulley et al., Phase III Trial of PROSTVAC in Asymptomatic or Minimally Symptomatic Metastatic Castration-Resistant Prostate Cancer. Journal of clinical oncology : official journal of the American Society of Clinical Oncology 37, 1051–1061 (2019).

6. P. J. Southall et al., Immunohistological distribution of 5T4 antigen in normal and malignant tissues. British Journal of Cancer 61, 89–95 (1990).

7. K. J. Ewer et al., Protective CD8+ T-cell immunity to human malaria induced by chimpanzee adenovirus-MVA immunisation. Nat Commun 4, 2836 (2013).

8. R. D. Antrobus et al., Clinical assessment of a novel recombinant simian adenovirus ChAdOx1 as a vectored vaccine expressing conserved Influenza A antigens. Molecular therapy : the journal of the American Society of Gene Therapy 22, 668–674 (2014).

9. N. Borthwick et al., Vaccine-elicited human T cells recognizing conserved protein regions inhibit HIV-1. Molecular therapy : the journal of the American Society of Gene Therapy 22, 464–475 (2014).

10. L. Swadling et al., A human vaccine strategy based on chimpanzee adenoviral and MVA vectors that primes, boosts, and sustains functional HCV-specific T cell memory. Science translational medicine 6, 261ra153 (2014).

11. K. Ewer et al., A Monovalent Chimpanzee Adenovirus Ebola Vaccine Boosted with MVA. The New England journal of medicine 374, 1635–1646 (2016).

12. A. B. Tiono et al., First field efficacy trial of the ChAd63 MVA ME-TRAP vectored malaria vaccine candidate in 5-17 months old infants and children. PloS one 13, e0208328 (2018).

13. N. Venkatraman et al., Safety and Immunogenicity of a Heterologous Prime-Boost Ebola Virus Vaccine Regimen in Healthy Adults in the United Kingdom and Senegal. The Journal of infectious diseases 219, 1187–1197 (2019).

14. F. Cappuccini, S. Stribbling, E. Pollock, A. V. Hill, I. Redchenko, Immunogenicity and efficacy of the novel cancer vaccine based on simian adenovirus and MVA vectors alone and in combination with PD-1 mAb in a mouse model of prostate cancer. Cancer immunology, immunotherapy : CII 65, 701–713 (2016).

15. F. Cappuccini, E. Pollock, S. Stribbling, A. V. S. Hill, I. Redchenko, 5T4 oncofoetal glycoprotein: an old target for a novel prostate cancer immunotherapy. Oncotarget, (2017).

16. S. J. McConkey et al., Enhanced T-cell immunogenicity of plasmid DNA vaccines boosted by recombinant modified vaccinia virus Ankara in humans. Nat Med 9, 729–735 (2003).

17. M. O. Afolabi et al., A phase I randomized clinical trial of candidate human immunodeficiency virus type 1 vaccine MVA.HIVA administered to Gambian infants. PloS one 8, e78289 (2013).

18. R. Harrop et al., MVA-5T4-induced immune responses are an early marker of efficacy in renal cancer patients. Cancer immunology, immunotherapy : CII 60, 829–837 (2011).

19. P. S. Gill et al., The handling and sampling of radical prostatectomy specimens for reporting and research: the Oxford approach. J Clin Pathol 65, 1057–1061 (2012).

20. N. Harder et al., Tissue Phenomics for prognostic biomarker discovery in low- and intermediate-risk prostate cancer. Sci Rep 8, 4470 (2018).

21. P. F. Schellhammer et al., Lower baseline prostate-specific antigen is associated with a greater overall survival benefit from sipuleucel-T in the Immunotherapy for Prostate Adenocarcinoma Treatment (IMPACT) trial. Urology 81, 1297–1302 (2013).

22. D. N. Amato RJ, Naylor S et al. ,Vaccination of Prostate Cancer Patients With Modified Vaccinia Ankara Delivering the Tumour Antigen 5T4 (TroVax): A Phase 2 Trial. Journal of immunotherapy 31, 577–585 (2008).

23. L. Coughlan et al., Heterologous Two-Dose Vaccination with Simian Adenovirus and Poxvirus Vectors Elicits Long-Lasting Cellular Immunity to Influenza Virus A in Healthy Adults. EBioMedicine 29, 146–154 (2018).

24. D. G. McNeel et al., Safety and immunological efficacy of a DNA vaccine encoding prostatic acid phosphatase in patients with stage D0 prostate cancer. Journal of clinical oncology : official journal of the American Society of Clinical Oncology 27, 4047–4054 (2009).

25. J. T. Becker et al., DNA vaccine encoding prostatic acid phosphatase (PAP) elicits long-term T-cell responses in patients with recurrent prostate cancer. Journal of immunotherapy 33, 639–647 (2010).

26. D. G. McNeel et al., Concurrent, but not sequential, PD-1 blockade with a DNA vaccine elicits anti-tumor responses in patients with metastatic, castration-resistant prostate cancer. Oncotarget 9, 25586–25596 (2018).

27. H. Kubler et al., Self-adjuvanted mRNA vaccination in advanced prostate cancer patients: a first-in-man phase I/IIa study. J Immunother Cancer 3, 26 (2015).

28. D. T. Le, E. M. Jaffee, Regulatory T-cell modulation using cyclophosphamide in vaccine approaches: a current perspective. Cancer research 72, 3439–3444 (2012).

29. M. Scurr et al., Effect of Modified Vaccinia Ankara–5T4 and Low-Dose Cyclophosphamide on Antitumor Immunity in Metastatic Colorectal Cancer: A Randomized Clinical Trial. JAMA Oncology, (2017).

30. L. Fong et al., Activated lymphocyte recruitment into the tumor microenvironment following preoperative sipuleucel-T for localized prostate cancer. J Natl Cancer Inst 106, (2014).

31. B. Farsaci et al., Identification by digital immunohistochemistry of intratumoral changes of immune infiltrates after vaccine in the absence of modifications of PBMC immune cell subsets. International journal of cancer. Journal international du cancer 135, 862–870 (2014).

32. M. Mercader et al., T cell infiltration of the prostate induced by androgen withdrawal in patients with prostate cancer. Proc Natl Acad Sci U S A 98, 14565–14570 (2001).

33. A. Strasner, M. Karin, Immune Infiltration and Prostate Cancer. Front Oncol 5, 128 (2015).

34. S. Vesalainen, P. Lipponen, M. Talja, K. Syrjanen, Histological grade, perineural infiltration, tumour-infiltrating lymphocytes and apoptosis as determinants of long- term prognosis in prostatic adenocarcinoma. Eur J Cancer 30A, 1797–1803 (1994).

35. J. Irani et al., High-grade inflammation in prostate cancer as a prognostic factor for biochemical recurrence after radical prostatectomy. Pathologist Multi Center Study Group. Urology 54, 467–472 (1999).

36. P. A. McArdle et al., The relationship between T-lymphocyte subset infiltration and survival in patients with prostate cancer. Br J Cancer 91, 541–543 (2004).

37. V. Karja et al., Tumour-infiltrating lymphocytes: A prognostic factor of PSA-free survival in patients with local prostate carcinoma treated by radical prostatectomy. Anticancer Res 25, 4435–4438 (2005).

38. S. G. Zhao et al., The Immune Landscape of Prostate Cancer and Nomination of PD- L2 as a Potential Therapeutic Target. J Natl Cancer Inst 111, 301–310 (2019).

39. M. A. McNamara, D. J. George, Pain, PSA flare, and bone scan response in a patient with metastatic castration-resistant prostate cancer treated with radium-223, a case report. BMC cancer 15, 371 (2015).

40. F. H. Karzai, R. A. Madan, W. D. Figg, Beyond PSA: managing modern therapeutic options in metastatic castration-resistant prostate cancer. South Med J 108, 224–228 (2015).

41. Y. Yamamoto et al., Tumour and immune cell dynamics explain the PSA bounce after prostate cancer brachytherapy. Br J Cancer 115, 195–202 (2016).

42. F. S. Hodi et al., Immune-Modified Response Evaluation Criteria In Solid Tumors (imRECIST): Refining Guidelines to Assess the Clinical Benefit of Cancer Immunotherapy. Journal of clinical oncology : official journal of the American Society of Clinical Oncology 36, 850–858 (2018).

43. K. Kim Bines and Kaufman, Trovax, a recombinant modified vaccinia ankara virus encoding 5T4. Lessons learned and future development. Human Vaccines 6, 784–791 (2010).

44. L. T. Nguyen et al., Expansion and characterization of human melanoma tumor-infiltrating lymphocytes (TILs). PloS one 5, e13940 (2010).

45. N. Brieu, O. Pauly, J. Zimmermann, G. Binnig, G. Schmidt, Slide-specific models for segmentation of differently stained digital histopathology whole slide images. SPIE Medical Imaging (SPIE, 2016), vol. 9784.

46. A. Kapil et al., Deep Semi Supervised Generative Learning for Automated Tumor Proportion Scoring on NSCLC Tissue Needle Biopsies. Sci Rep 8, 17343 (2018).

